# Evolution of Chronic Lesion Tissue in RRMS patients: An association with disease progression

**DOI:** 10.1101/2023.12.29.23300651

**Authors:** Samuel Klistorner, Michael H Barnett, John Parratt, Con Yiannikas, Alexander Klistorner

## Abstract

**Background and Objective:** This study examines the long-term changes in Chronic Lesion Tissue (CLT) among relapsing and remitting MS (RRMS) patients, focusing on its impact on clinical and radiological disease progression indicators.

**Methods:** The study involved 72 MS patients with at least a 5-year follow-up. Annual assessments used 3D FLAIR, pre- and post-contrast 3D T1, and diffusion-weighted MRI. Lesion segmentation was conducted using iQ-MS^TM^ software, while brain structures were segmented using AssemblyNet. Volumetric changes in CLT were tracked using a custom-designed pipeline.

**Results:** Throughout the follow-up period, the volume of CLT in the entire cohort increased continuously and steadily, averaging 7.75±8.2% or 315±465 mm³ per year. Patients with expanding CLT experienced significantly faster brain atrophy, affecting both white and grey matter, particularly in the brain’s central area. Expanded CLT was also associated with higher and worsening EDSS scores, in contrast to the stable CLT group, where EDSS remained unchanged.

**Conclusion:** This study demonstrates that, over a period of up to 7 years, patient-specific enlargement of CLT, where present, progresses at a constant rate and significantly influences disease progression.

## Introduction

Multiple sclerosis (MS) is a complex central nervous system (CNS) disorder characterised by heterogenous inflammatory and variably destructive white and grey matter lesions. Smouldering inflammation at the rim of a subset of chronic lesions not only signifies the progressive nature of MS but also plays an instrumental role in disability worsening in patients with established disease.[1] This low-grade inflammatory activity and demyelination manifests as a slow expansion of chronic lesion tissue (CLT) over time, eventually leading to axonal transection, neurodegeneration and disability progression.[2][3][4][5][6]

Despite the critical role that CLT enlargement plays in the MS narrative, some aspects of its longitudinal evolution are not fully understood. Questions regarding its duration, consistency over time, or patient-specific nature have not been adequately examined. Addressing these knowledge gaps could significantly improve our understanding of smouldering lesions, help refine patient selection criteria for future clinical trials of agents that putatively target chronic inflammation and, ultimately, enhance our ability to treat and manage MS.

However, substantial variations exist in the methodologies employed to identify and monitor the enlargement of chronic lesions. These variations pertain to how chronic expanding lesions are selected, specifically the threshold used to assign lesions to the slowly expanding lesion (SEL) category. Additionally, variations are observed in how these lesions are measured, whether by counting the number of expanding lesions or by measuring the volume of chronic lesion enlargement. For example, the widely used Jacobean transformation-based technique is useful for identifying and counting individual lesions that meet specific classification criteria for SELs. However, this method is not suitable for the longitudinal volumetric estimation of lesion expansion. On the other hand, assessing chronic lesion expansion through manual lesion segmentation, although likely more accurate in estimating the actual volume of expanded lesional tissue, is time-consuming and may have limited applicability in large cohorts or serial studies.

Therefore, we introduced a novel pipeline that estimates longitudinal volumetric changes in chronic lesion tissue using serial MRI data, integrating Artificial Intelligence (AI)-assisted lesion segmentation. This algorithm extends our previous technique, transitioning from a two-timepoint approach to a multiple-point analysis. The method not only accounts for lesion shift due to brain atrophy between timepoints, but also identifies and removes new and confluent lesions (which are presumably driven by conventional, acute inflammatory demyelination) that emerge during the study. Importantly, it eliminates the need for chronic lesion classification.

In this study, we applied this novel approach to investigate the long-term evolution of chronic lesion tissue and its impact on clinical and radiological measures of disease progression. We conducted this analysis using data obtained from a cohort of Relapsing-Remitting Multiple Sclerosis (RRMS) patients who were followed for up to 7 years. Additionally, we developed an imaging protocol and calculated the required sample size for trials involving therapeutic agents aimed at reducing smouldering inflammation.

## Methods

### Subjects

Consecutive patients with established RRMS who were enrolled in the MADMS study (National MS Society grant, G-1508-05946) and had reached 51years of follow-up were included. MS was diagnosed according to the revised McDonald 2010 criteria. Patients underwent annual MRI scans and clinical assessment. Patients were relapse-free and their therapy did not change for at least 61months prior to enrolment. In addition, no steroid treatment was administered for at least 61months prior to enrolment.

Disease Modifying Therapy (DMT) was categorised into 3 groups: no treatment, low efficacy treatment (injectables, such as interferon and glatiramer acetate, teriflunomide and dimethyl fumarate) and high-efficacy treatment (fingolimod, natalizumab, alemtuzumab and ocrelizumab). [7]

### MRI protocol and analysis

MRI was performed using a 3T GE Discovery MR750 scanner (GE Medical Systems, Milwaukee, WI). The following MRI sequences were acquired: Pre- and post-contrast (gadolinium) Sagittal 3D T1, FLAIR CUBE, diffusion-weighted MRI. Specific acquisition parameters and MRI image processing are presented in the Supplementary material.

### Lesion analysis

White matter lesions were segmented for each time point of each patient using an AI-based lesion segmentation algorithm (iQ-Solutions^TM^-MS Report, hereafter referred to as iQ-MS^TM^ software suite, Sydney Neuroimaging Analysis Centre, Sydney, Australia).[8] The segmentation was based on unprocessed T1 and FLAIR images. These lesion masks were then transformed to ACPC space using linear registration. Only lesions larger than 50mm³ were included in the analysis.

To verify the AI algorithm’s ability to correctly identify and measure the longitudinal change of chronic lesion volume, manual lesion segmentation was also implemented in a subgroup of 25 patients, and the gradient of lesion expansion between automated and manual modes was compared. Manual segmentation was performed using JIM 9 software (Xinapse Systems, Essex, UK) on co-registered T2 FLAIR images.

Due to continuous change in the morphology and intensity of individual lesions, which may result in the merging or separation of adjacent lesions on sequential scans, it is frequently impractical to track a single lesion over long-term serial MRIs. [9][10] Therefore, in the current study, our focus was not on tracking the expansion of individual lesions but rather on estimating the total change in chronic lesion tissue volume at the patient level. To achieve this, we made significant modifications to the previously described Lesion Estimation and Analysis Pipeline.[5]

The main building block of the new pipeline was a pair of co-registered lesion masks from two sequential MRIs. The pipeline implemented the following steps:

1. Matching of individual lesions between MRIs obtained at timepoints 1 and 2,
2. Classification of all timepoint 2 lesions without a matched counterpart at timepoint 1 as ‘new free-standing lesions,’ and excluding them from CLT analysis,
3. Spatial adjustment of remained lesions to account for brain atrophy through affine co-registration between the two masks, as described in [5],
4. Identification of new confluent lesions at timepoint 2 using a custom-designed algorithm that detects non-spherical areas of lesion expansion; and excluding them from CLT analysis, as described in [5],
5. Classification of gadolinium-enhancing lesions and lesions undergoing substantial shrinking as ‘acute’ or ‘post-acute lesions’ respectively; and excluding them from CLT analysis (detailed description provided in Supplementary material),
6. Calculation of the volume of all remaining lesion tissue for timepoints 1 and 2, with the difference specified as ‘annual chronic lesion tissue change.’

Subsequently, a cumulative volume (or %-wise increase) of CLT expansion for each patient is computed by summing consecutive annual expansion values. This process constructs a patient-wise profile of CLT evolution (Fig. 1) based on the line of best fit.

**Fig. 1.**
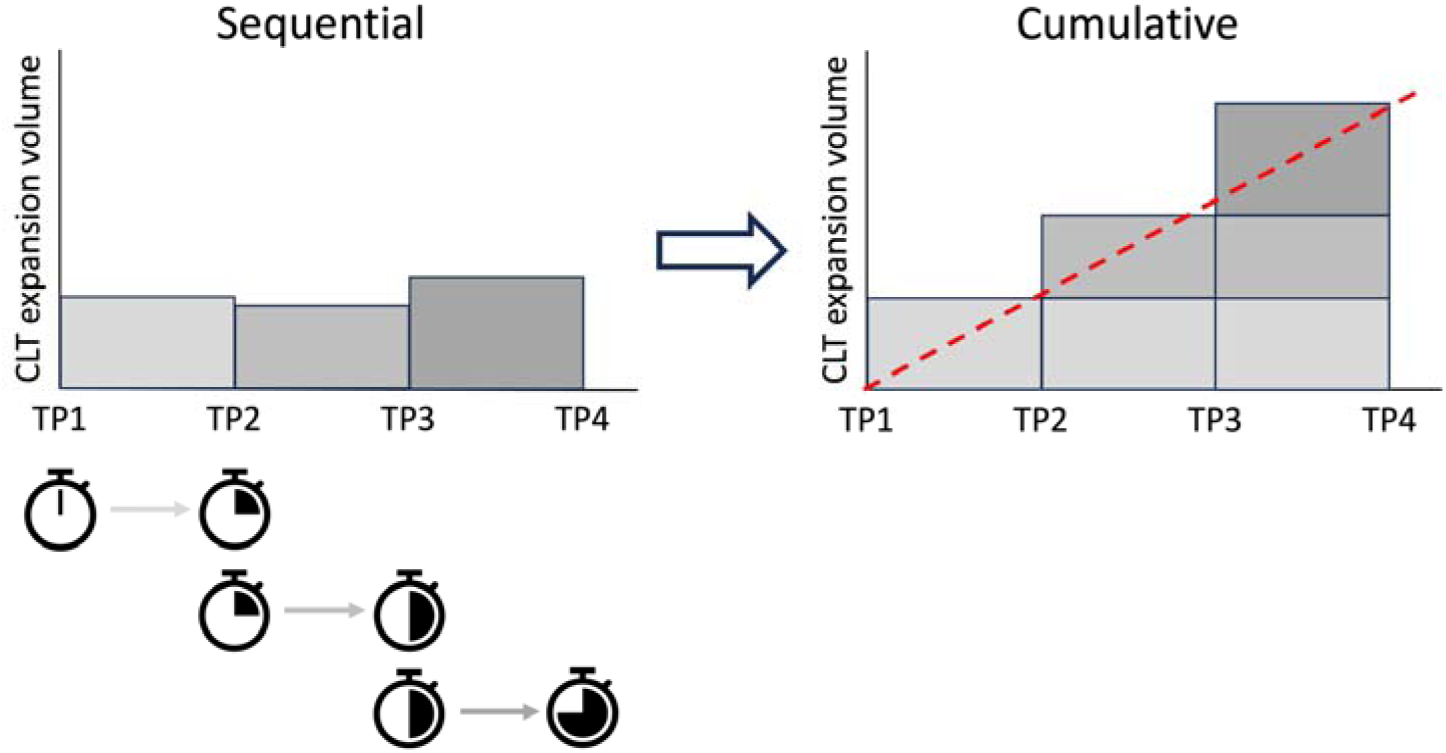
Calculation of cumulative volume of chronic lesion tissue (CLT) expansion. CLT change is measured between two sequential scans for each annual interval independently. The cumulative profile is calculated by summing consecutive annual CLT expansion values.

The degree of tissue damage within chronic lesions was assessed by mean diffusivity (MD), as previously proposed.[11] Progressive annual tissue damage in chronic lesions was measured as an increase of MD in the lesion core between two consecutive MRI scans. A cumulative MD change for each patient was computed by adding the annual change.

### Volumetric brain analysis

Volumetric measures were performed using AssemblyNet, an AI brain segmentation tool, [12] on T1 images co-registered to ACPC space. The following volume metrics were analysed: total brain, white matter, grey matter, cortex and deep grey matter and ventricular volume.[13] [14] The cerebellum was excluded from the white and grey matter analysis. All volumes were scaled based on MNI template registration to account for head size variability.

For longitudinal analysis, to minimise the impact of variation in segmentation, we used “a volume change per year” metric as a measure of atrophy. This involved calculating the annual change for each MRI measure and generating a line of best fit to represent the annual changes. The gradient of the line of best fit represented the annual volume change.

### Statistics

Continuous variables were assessed for normality using the Shapiro–Wilk normality test and described as mean and standard deviation (SD) or median with interquartile range (IQR), as appropriate.

The chi-square statistic was used for sex comparison between groups. For Expanded Disability Status Scale (EDSS) and Disease Modifying Therapy (DMT) at baseline Independent-Samples Mann-Whitney U Test was used to evaluate significant difference between groups.

For longitudinal changes of EDSS Related-Samples Wilcoxon Signed Rank Test was used. Significance of age and disease duration between groups was determined using Students’ t-test.

Analysis of Covariance (ANCOVA) was used to determine the significant difference of brain atrophy measures between groups. Baseline analysis was controlled for age and sex, while longitudinal analysis was also controlled for new lesion volume.

p < 0.05 was considered statistically significant.

## Results

Seventy-two consecutive MS patients enrolled in a longitudinal study of MS, who reached 5 years of follow-up, were included in this study. Of these, 17 patients reached 7 years and were included in the sub-analysis. Four subjects were removed from the analysis due to the very low (<100mm^3^ ) volume of chronic lesions. Demographic data presented in Table 1

**Table 1.** Demographic data.

|  | 5 years follow-up group | 7 years follow up group |
| --- | --- | --- |
| Total subjects | 68 | 17 |
| Age at enrolment, y | 40.9 ± 9.8 | 42.4 ± 9.9 |
| Gender | 25M 43F | 7M 10F |
| Disease duration at enrolment, y | 6.2 ± 5.1 | 5.6 ± 3.4 |
| EDSS at baseline, median (range) | 1 (0-3) | 1 (0-3) |
| Lesion volume at baseline | 4193 ± 4669 | 4039.0 ± 4025.8 |

At the start of the study, 23 patients were on low-efficacy treatment (injectables, such as interferon and glatiramer acetate, teriflunomide and dimethyl-fumarate), 38 patients were receiving high-efficacy drugs (fingolimod, natalizumab, alemtuzumab, ocrelizumab) and seven patients were treatment-free. During the study, 20 patients changed the treatment category. By the end of the study period, 12 patients remained on low-efficacy treatment and 48 patients were on high-efficacy drugs. The decision to modify the therapy was taken by the treating physician based on combination of clinical and radiological activity. Overall, 48 patients did not change treatment category during the study.

### Manual vs IQMS segmentation comparison

We observed a strong correlation between the gradients of CLT expansion obtained using manual and AI-based lesion segmentation (r = 0.96, p < 0.001, Suppl Fig.1). This confirms the suitability of IQ-MS^TM^ AI-based segmentation for our dataset and in-house pipeline, and we proceeded with IQMS segmentation for the remaining analysis.’

### Progressive CLT enlargement is constant during 7 years of follow-up

Upon visual examination of CLT enlargement profiles of individual patients, it became evident that subject-wise expansion is a relatively constant process, typically well approximated by a linear function. Examples of CLT expansion profiles in individual patients are presented in Fig. 2. To quantitatively assess the predictive accuracy of the linear model, we calculated the Root Mean Square Error (RMSE) of cumulative CLT enlargement rate for each subject. The RMSE values were generally low for most patients, with only three exceptions (median 4.9, interquartile range 3.3-7.1), supporting our observation (Suppl Fig.2). Therefore, the magnitude of CLT expansion for individual subjects can be effectively described by the gradient of the linear function representing the degree of absolute (volumetric) or relative (%) CLT expansion per year. This gradient was subsequently used in further analyses, including comparisons with clinical and radiological data.

**Fig. 2.**
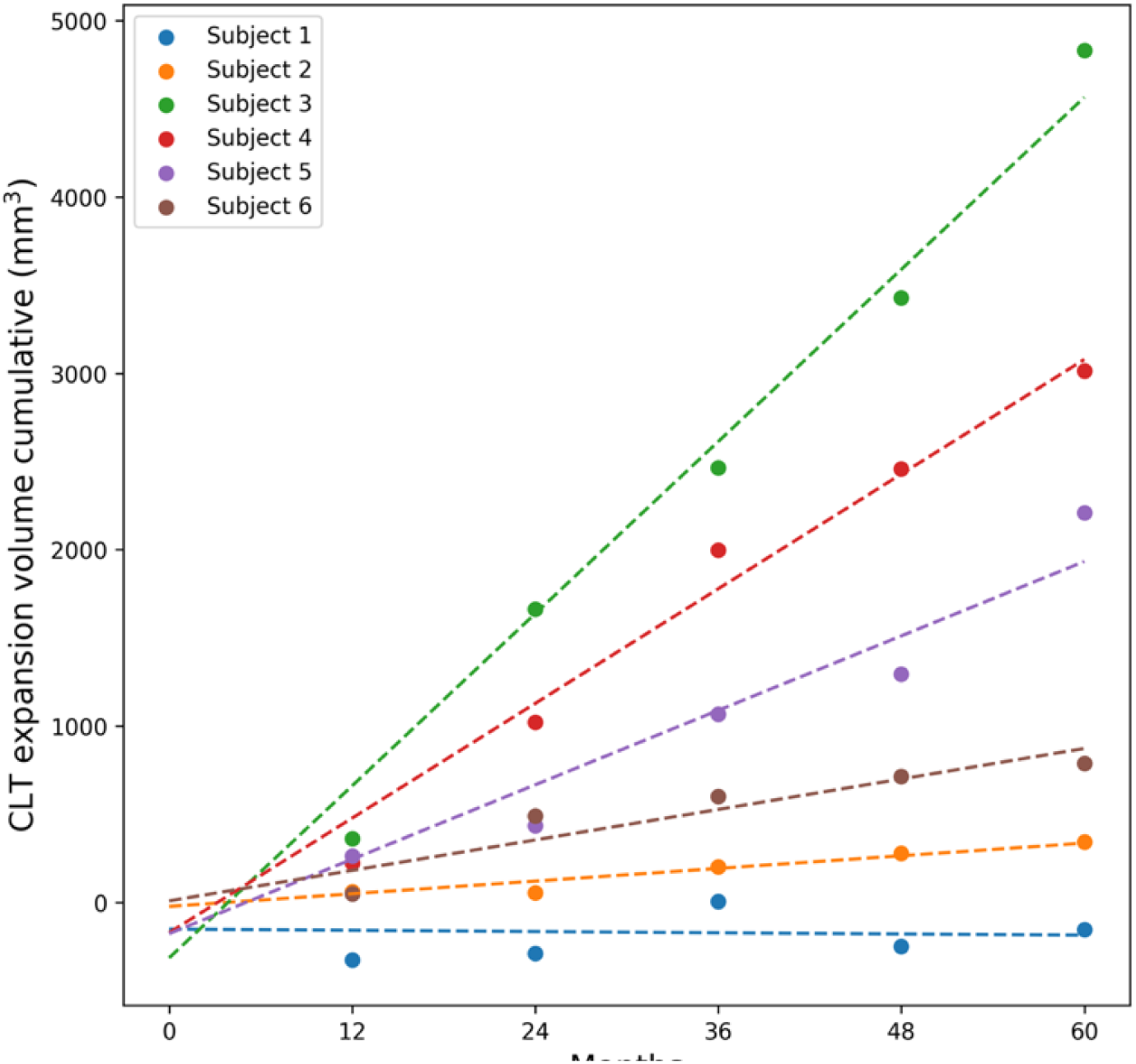
Examples of CLT expansion profiles in individual patients. Dotted lines represent best linear fit.

The average cumulative rate (%) of CLT enlargement in the entire cohort is depicted in Fig. 3a. The figure illustrates a continuous and steady increase in the relative volume of CLT throughout the entire follow-up period, with an average annual increase of 7.75+/-8.2%. A similar linear trend was observed in the sub-group of patients who reached 84 months of follow-up (Fig. 3b).

**Fig. 3.**
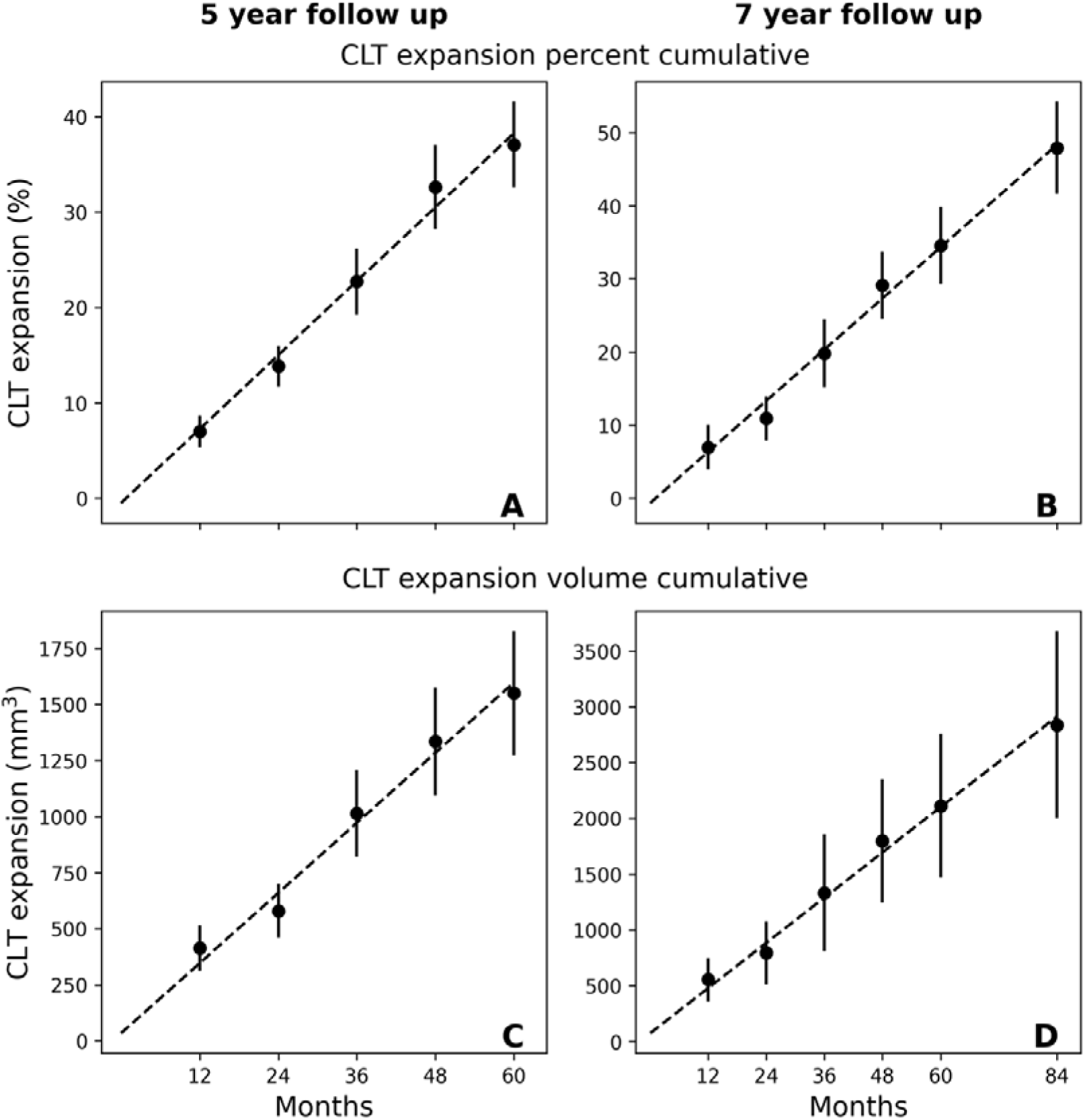
(a-d). CLT expansion in entire patients cohort (a, c) and sub-group of patients who reached 7 years follow-up. Upper row represents a relative (%-wise) change of chronic lesion volume, lower row shows an absolute values of CLT change.

In cases with small baseline lesion volumes, however, even a significant percentage of CLT expansion is unlikely to result in clinically or radiologically meaningful progression. Therefore, to estimate the impact of CLT enlargement on disease evolution, we also examined the annual increase in the absolute volume of expansion. This analysis revealed a similar linear growth pattern for the entire cohort over 60 months of follow-up, with an average expansion of approximately 315+/-465 mm³ per year (Fig. 3c). The linear nature of CLT increase was also observed in a smaller sub-group of patients who reached 7 years of follow-up (Fig. 3d).

The gradient of CLT volume enlargement exhibited significant variation among individual patients, spanning from around zero to over 2400 mm²/year. Ranking patients based on the values of patient-wise CLT gradients revealed a continuum of expansion magnitudes between individual subjects, making it difficult to conclusively dichotomize them into CLT expanding or CLT stable groups (Fig. 4a).

**Fig. 4 (a, b).**
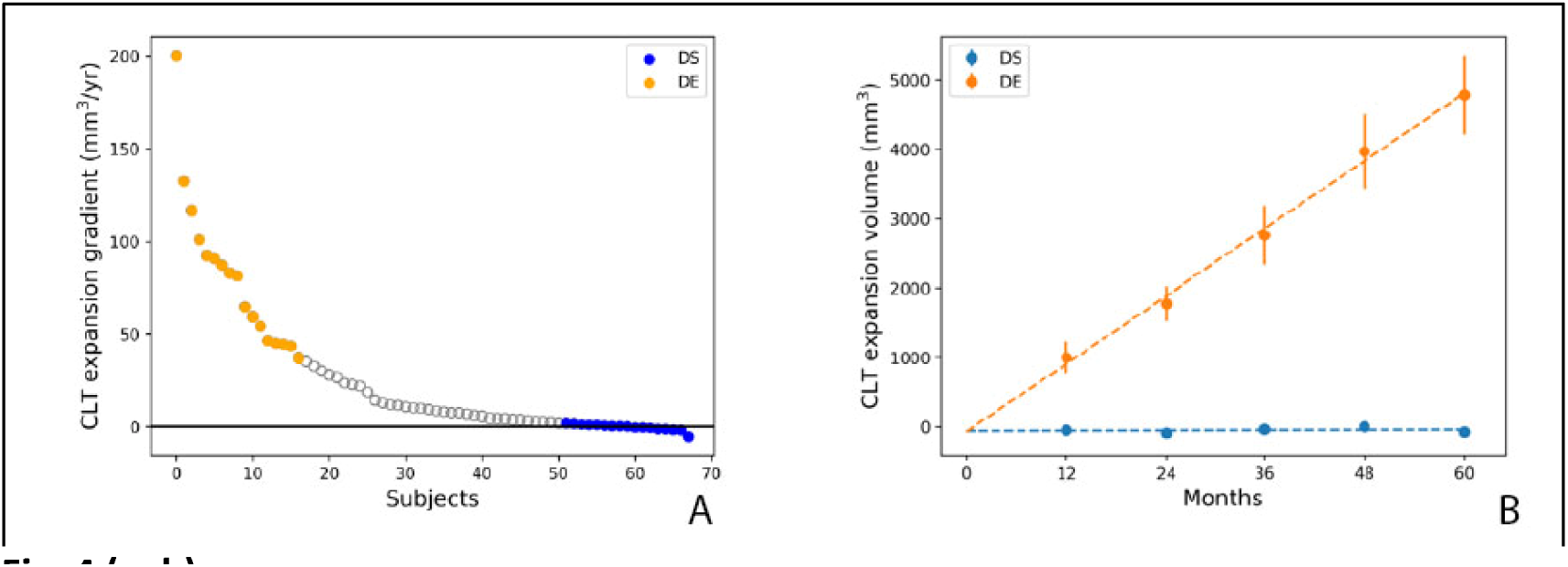
a. Ranking of CLT gradients. Orange circles indicate the top quartile (“Definitely Enlarging” group), blue circles indicate the bottom quartile (“Definitely Stable” group) b. Annual CLT expansion in “Definitely Enlarging” and “Definitely Stable” groups

Therefore, to examine the effect of chronic lesion activity on clinical and radiological measures of disease progression, we divided patients into quartiles based on the extent of CLT enlargement. The top quartile (orange) and the bottom quartile (blue) were designated as categorical extremes, forming the Definitely Expanding group (DE) and the Definitely Stable group (DS), respectively, with each group consisting of 17 subjects. The effectiveness of this separation is evident in Fig. 4b, which illustrates a substantial and consistent annual increase in CLT volume in the DE group (970+/-115 mm3), while showing no visible volume change in the DS group.

Significant differences in the degree of CLT enlargement between the two groups became evident by the second year of the follow-up (year 1: p = 0.19, year 2: p < 0.001, t-test), a critical factor in calculating the sample size for a potential clinical trial (see below).

### CLT volume enlargement is associated with loss of (predominantly central) brain tissue and EDSS progression

There was a statistically significant difference between the groups in terms of age (p=0.04, t-test) and gender composition (3 times more males in the expanding group, p=0.03, Fisher exact test). Consequently, all presented brain volumetric comparisons at baseline were adjusted for age and sex.

Baseline brain volumetric data for the DE and DS groups are presented in Table 2 and Figure 5 (left column). Group-based analysis revealed a significantly smaller total brain volume, as well as white, grey and deep grey matter volume, in the DE group compared to the DS group. In contrast, total and cortical grey matter did not differ between the two groups. The difference in ventricle volumes demonstrated borderline significance.

**Fig. 5.**
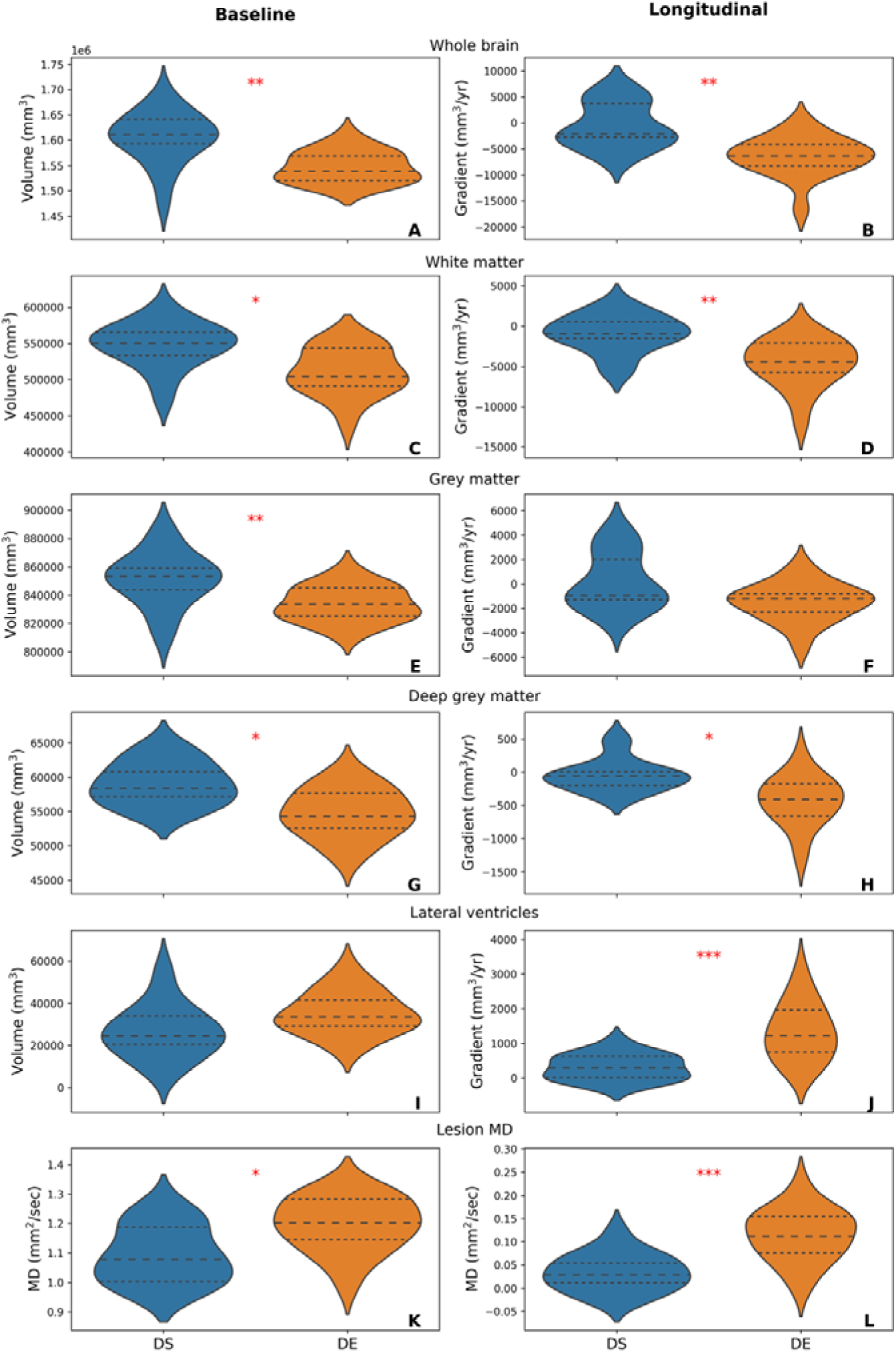

**Table 2.**
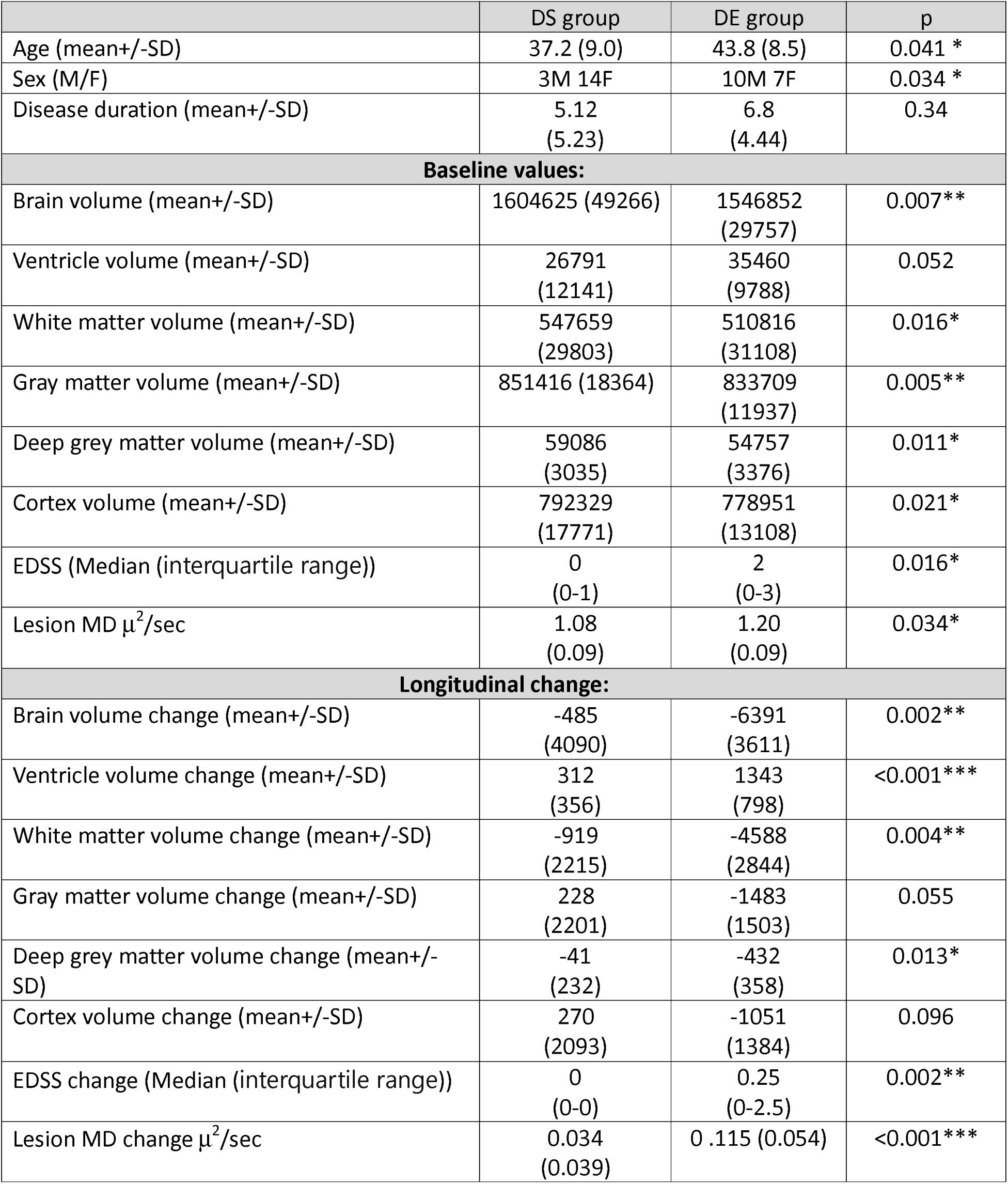
Comparison of baseline and longitudinal brain volumetric data between the DE and DS groups.

The composition of disease-modifying therapies (DMT) was not significantly different between the two groups at baseline (p = 0.074, Mann-Whitney U Test).

Longitudinal analysis of brain volumetrics revealed significantly faster brain atrophy in the DE group compared to the DS group during the follow-up period (see Table. 2 and Fig. 5 b).

This atrophy affected both total white and total grey matter (Fig. 5d) and was particularly pronounced in the central area of the brain, impacting both white matter (central brain atrophy, measured by an increase in ventricular size, Fig. 5 j) and grey matter (deep grey matter atrophy, Fig. 5 h). However, no significant difference was observed between the groups in terms of cortical thinning. To account for the significantly larger volume of new lesions in the DE group on brain volumetrics, this parameter was included as a covariate in the longitudinal model.

In the DE group, CLT exhibited a significantly higher MD value at baseline compared to the DS group (1.20+/-0.09 and 1.08+/-0.09, respectively, p=0.034). Additionally, the average increase in MD was significantly larger in the DE group than in the DS group (0.115+/-0.054 and 0.034+/-0.039, respectively, p<0.001). This indicates more extensive tissue damage within chronic lesions in the expanding group, both at baseline and during follow-up.

EDSS was also significantly higher in the DE group at baseline (mean 1.6, median 2 vs. mean 0.43, median 0, p=0.016, Independent-Samples Mann-Whitney U Test). During the study, EDSS significantly worsened in the expanding group (1.6 vs. 2.5, p=0.04, Related-Samples Wilcoxon Signed Rank Test), while remaining unchanged in the stable group (0.43 vs. 0.40), resulting in a more significant EDSS difference at the last follow-up (mean 2.5, median 2.5 vs mean 0.40 median 0, p=0.002, Independent-Samples Mann-Whitney U Test).

### Sample size calculation for a clinical trial investigating the effect of treatment on slow expansion of chronic lesions

The sample size calculation for a hypothetical placebo controlled clinical trial to test the effect of treatment on slow expansion of chronic lesions was based on the average annual slope and standard deviation (SD) of CLT expansion in the DE patient group. Considering that a reduction in the lesion expansion slope can occur only in cases with an established pattern of lesion expansion, it is essential to select only patients who have demonstrated a significant gradient of Chronic Lesion Tissue (CLT) expansion before the trial period. Based on our analysis, such patients constituted one quartile of the entire cohort.

To identify patients suitable for such a trial, the MRI results from the two years preceding enrolment must be examined (since, as shown above, 2 years is sufficient to observe a statistical difference between two groups). A minimum of three pre-trial MRI time-points are needed to determine the expanding gradient. Since only a quarter of RRMS population with the highest expansion gradient is suitable for the study, MRI scans of a group four times the size of the required enrolment number must be screened. Inclusion also should be limited to patients whose pre-trial scans were acquired with the same scanner/imaging protocol.

Reduction of the CLT enlargement slope in the treated group is indicative of the potential ‘treatment effect’, as shown in Fig. 6a (arrow). Fig. 6b illustrates the sample size (per arm) required for a treatment effect ranging between 20% and 60%, which is best described by power function. For instance, to demonstrate a statistically significant (two-sided, p<0.05) reduction in lesion expansion with 80% power for a treatment effect of 50%, it would be sufficient to enrol 24 patients per arm. Consequently, to meet this requirement, the pre-trial MRIs of (24×4×2 arms) = 192 patients would need to be screened.

**Fig. 6.**
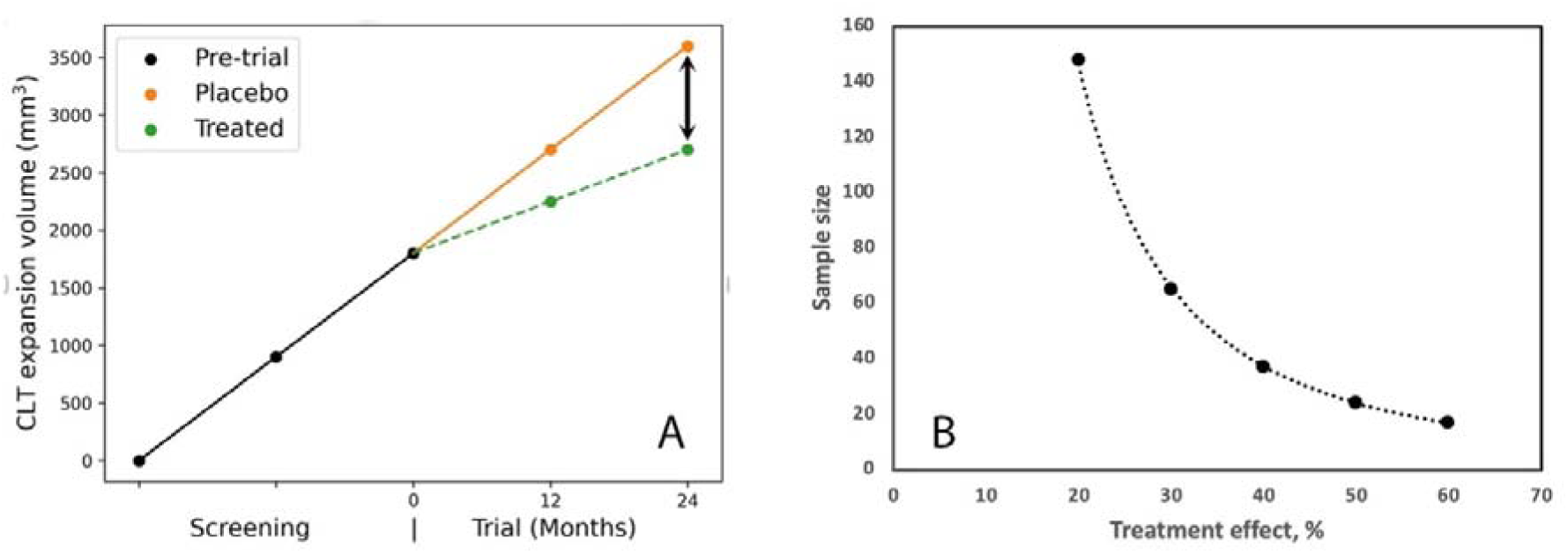

## Discussion

This study investigated the long-term longitudinal dynamics of CLT expansion in patients with RRMS and its association with clinical and radiological biomarkers of disease progression. We established that the slow expansion of chronic lesion tissue is a patient-specific process, which, when present, remains remarkably constant, at least within the timeframe of this study (i.e., up to 7 years). Furthermore, we demonstrated a significant link between CLT expansion and clinically and radiologically measured disease progression. Additionally, we observed a relatively higher prevalence of this expansion among male patients.

These observations were facilitated by the relatively long study duration and the consistent use of the same imaging hardware and protocol. There are also two novel technical aspects of the study that enabled us to accurately estimate subtle longitudinal change in CLT volume. Namely, the utilisation of AI-based lesion segmentation and the development of a new serial MRI pipeline for the identification and measurement of the volume of the slowly expanding component of chronic lesions.

The potential benefits of AI, such as speed and consistency, make it an attractive alternative to manual or semi-automated techniques for estimating lesion expansion. Given that applying AI for this purpose is a novel approach, we sought to validate its accuracy and reliability. To achieve this, we compared the CLT expansion gradient between AI-based lesion segmentation and a thorough manual segmentation method in a representative patient’s sub-group. This comparison revealed a very high level of concordance, confirming the suitability of using the AI-based technique for the entire cohort.

The new pipeline offers several advantages for studying the expansion of chronic lesion tissue over multiple serial MRIs. In contrast with our previous methodology[5][15][16], which was limited to the investigation of chronic lesion expansion between two MRI scans taken several years apart, the current pipeline permits the incorporation of an extended series of MRI examinations.

The dynamic nature of this pipeline also enables the integration of newly occurring lesions into the analysis throughout the study. Owing to its pair-based annual approach for analyzing expansion between two consecutive scans, newly formed lesions can be accurately identified at each annual interval. These lesions are then included in the analysis from the time they become chronic, namely at least 12 months after their initial appearance.

Furthermore, since our analysis measures the entire volume of chronic lesion tissue expansion for the subject, rather than the number of expanding lesions, it eliminates the need for chronic lesion classification into slowly expanding lesions (SEL) and non-slowly expanding lesions (non-SEL) categories. Rather than requiring individual lesions to demonstrate ‘constant’ enlargement across at least three time-points based on heuristically determined thresholds, the current algorithm directly estimates the actual volume of expanded chronic lesion tissue in individual patients. However, we still appled a criterion for ‘asphericity’ of lesion growth to identify (and exclude) new confluent lesion activity..

### The Constant and Patient-Specific Nature of CLT Expansion

Using the new pipeline, we established that, on average, the volume of CLT in our cohort of RRMS patients increased gradually and constantly over the course of the study, i.e., up to 7 years. We also demonstrated that the gradient of CLT expansion varied widely between patients, ranging from stable to extensively expanding.

However, in individual patients, the annual change of CLT enlargement remained relatively constant throughout the entire observation period (as supported by low RMSE values). This is also confirmed by the group analysis of patients who do experience CLT expansion, which demonstrates the remarkable stability of the expansion gradient. Therefore, CLT expansion appears to be a patient-specific process; expansion of chronic lesions does occur (to varying degrees) in some patients, while others do not experience it.

Currently, the reason why some patients experience the growth of chronic lesion tissue while others do not remains unclear. Numerous studies have linked smouldering inflammation at the rim of chronic lesions (and lesion expansion as its imaging equivalent) with microglial activation and low-grade inflammatory demyelination.[17][3],[18][18][19] In light of this, recent publications suggesting that patient-specific factors are more likely to influence microglial response and oligodendroglial pathology in MS are particularly noteworthy. These individual factors may include age, sex, disease stage, lesion location, genetic heterogeneity, susceptibility to de/remyelination or even lifestyle or comorbid conditions.[20][21][15][22][3]

Indeed, our findings show a higher prevalence of males in the DE group, which might be a contributing factor to the CLT expansion and associated neurodegeneration (as suggested by our earlier studies [11] [23]). This aligns with the well-known observation that male patients with MS tend to have a more aggressive disease course. [26][24][25] Patients in the DE group were also significantly older compared to the stable group, and their immune response may, therefore, be shifted from an adaptive to a compartmentalised, innate phenotype (and associated microglial activation). [25]

Another patient-specific factor that may potentially influence the severity of smouldering inflammation at the lesion rim, and consequently, the rate of CLT expansion, is the type of treatment a patient receives. It has recently been demonstrated that natalizumab treatment decreases the prevalence of slowly expanding lesions in Secondary Progressive MS patients [26],[27] likely by reducing microglial activation at the rim of chronic lesions. [27] Ocrelizumab was also reported to reduce the relative volume of slowly expanding lesions (but, surprisingly, not other measures of smouldering inflammation, such as a decrease in T1 signal intensity and an increase in T1 hypointense lesion load) in patients with primary progressive MS [4]. Although our sample size is too small to reliably estimate the potential effect of individual therapies, the proportion of patients receiving first and second-line therapies at baseline was similar between the DE and DS groups. Furthermore, the majority (10/17) of patients in the DE group were already on high-efficacy therapy at the beginning of the study. In addition, a further 5 patients in this group changed treatment from a less effective option to a more effective one without any visible effect on the gradient of CLT expansion. However, this question remains open; and the potential effect of therapy would be best assessed in the context of a treatment-specific clinical trial.

### Expansion of chronic lesion tissue is linked to brain tissue loss, predominantly in the central regions, and to disease progression

Ranking patients based on the degree of CLT expansion did not reveal a clear dichotomy. Instead, it demonstrated a continuum of change in the expansion gradient.

Hence, to examine the effect of CLT expansion on disease progression, we employed quartiles rather than choosing a threshold to separate subjects into ‘expanding’ and ‘non-expanding’ categories. This approach identified the most expanding and most stable cases, designating them as the Definitely Expanding (DE) and Definitely Stable (DS) groups, respectively.

The longitudinal examination of CLT expansion in these groups affirmed the static nature of chronic lesion volume in the DS group and the ongoing growth in the DE group. Notably, in the DE group, the average cumulative rate of lesion expansion displayed a remarkably consistent linear trend across multiple time-points during the entire follow-up. Throughout this period, there was no evidence of either regression or acceleration of the annual expansion gradient.

Group analysis revealed a more prominent loss of brain tissue at baseline in the DE group. More importantly, there was also a faster rate of brain atrophy during the follow-up period in patients with definite expansion, compared to those in the stable group. The difference between the two groups remained highly significant after adjusting the analysis for the volume of new lesions, indicating a considerable contribution of CLT expansion to brain tissue loss. It is understood that smouldering inflammatory demyelination at the rim of chronic lesions leads to the transection of axons traversing the inflamed lesion rim. This results in both retrograde and anterograde (Wallerian) axonal degeneration, negatively impacting anatomically interconnected brain areas and contributing to accelerated brain atrophy.[28]

This trend was particularly evident in the central brain, as indicated by ventricular enlargement and deep grey matter atrophy, which showed the largest statistical differences between the groups. In contrast, the difference in longitudinal changes of cortical volume between the groups was non-significant. The prevalence of central brain tissue damage in patients with chronic lesion expansion is likely linked to the predominantly periventricular location of the expanding lesions.[15]

The link between lesion expansion and axonal damage is further strengthened by the more significant rarefaction of chronic lesion tissue (as measured by an increase in MD inside the lesions [5],[20]) in the DE group compared to stable patients. This difference was observed both at baseline and during follow-up. Elevated MD in chronic white matter MS lesions, reflects enlargement of the extracellular space,[29][30][31][32] which is caused by a combination of ongoing tissue loss and the relatively rigid structure of chronic lesion.[33] Therefore, the significantly higher MD at baseline and the larger increase in MD during follow-up observed in the DE group suggest that ongoing smouldering inflammation at the lesion edge, as evidenced by CLT expansion, promotes progressive axonal damage.

The debilitating effect of chronic lesion expansion on the progression of MS is further evidenced by observed differences in patient’s physical disability and its evolution over time. The DE group demonstrated a higher EDSS at baseline, which notably increased during the study period. In contrast, EDSS in the DS group remained stable. This disparity suggests that chronic lesion expansion, along with associated neurodegeneration, significantly contributes to physical decline in patients with MS. These findings are consistent with our previous work [5]. Moreover, the correlation between chronic lesion activity and physical disability has been similarly demonstrated in patients with primary progressive MS (PPMS)[4] and secondary progressive MS (SPMS).[26]

### Sample size calculation

In parallel with improved understanding of the role of smouldering inflammation (and associated lesion expansion) in disease progression, there is a growing interest in developing therapies to mitigate this process. We therefore performed a sample size estimation for a hypothetical clinical trial focusing on reducing the rate of chronic lesion expansion. Our calculations suggest that, to observe a reasonable treatment effect of 30-50%, a relatively small cohort of RRMS patients, ranging from 24 to 69 individuals per arm, would be required.

However, as only subjects with an established trend of lesion expansion are suitable for the trial, a thorough pre-trial screening process is necessary. This process involves the collection and analysis of multiple MRI compatible scans, which may not always be available.

### Study limitations

Our study has several limitations. First, our analysis was based on a relatively small sample of patients with RRMS, particularly when group analysis was involved. Larger studies are necessary to confirm these findings.

Secondly, while we acknowledge that different treatment options may exert varying effects on smouldering inflammation and associated lesion expansion, heterogeneity of treatment at baseline in our study and changes in medication during the follow-up period limit our ability to comprehensively assess their impact on our observations. Nevertheless, considering the consistent nature of the chronic lesion expansion process observed in this study, and the tendency for treatment alterations to involve a shift toward more potent drugs, it is likely that even currently available highly effective therapies, which primarily target the adaptive immune response, have a limited influence on this process.

Thirdly, no longitudinal analysis of individual lesions was performed in this study. However, due to temporal changes in lesion morphology in relation to merging of lesions or the development of new confluent lesions, the MRI time-series analysis implemented here is more appropriately suited for estimating lesion expansion on a patient-wise basis, rather than focusing on individual lesions. Importantly, MS therapy targets the patient, rather than specific, individual lesions.

In conclusion, our data show that, over a 7-year period, patient-specific enlargement of CLT, where present, evolves at a constant rate and significantly contributes to disease progression.

## Data Availability

All data produced in the present study are available upon reasonable request to the authors

