## Supplementary material for "Evolution of Chronic Lesion Tissue in RRMS patients: An association with disease progression"

### Supplementary methods

#### 1. Specific MRI parameters and image processing.

The following MRI sequences were acquired:

a. Pre- and post-contrast (gadolinium) Sagittal 3D T1: GE BRAVO sequence, duration 4 min each, FOV 256mm, Slice thickness 1mm, TE 2.7ms, TR 7.2ms, Flip angle 12°, Pixel spacing 1mm. Acquisition Matrix (Freq x Phase) is 256x256, which results in 1mm isotropic acquisition voxel size. The reconstruction matrix is 256x256.
b. FLAIR CUBE; GE CUBE T2 FLAIR sequence, duration 6 min, FOV 240mm, Slice thickness 1.2 mm, Acquisition Matrix (Freq x Phase) 256x244, TE 163ms, TR 8000ms, Flip angle 90°, Pixel spacing 0.47 mm. The reconstruction matrix is 512x512.
c. Echo-Planar Imaging based diffusion weighted MRI, duration 9 min (64-directions with 2 mm isotropic acquisition matrix, TR/TE = 8325/86 ms, b = 1000 s/mm$^2$, number of b0 = 2).

#### 2. MRI image pre-processing:

The baseline T1-weighted imaging was realigned to Anterior and Posterior Commissure (AC-PC) orientation. Using FLIRT (FSL, FMRIB Software Library), follow-up T1 images were co-registered to initial (month 0) AC-PC space by applying transformation matrices derived from linear co-registration between baseline AC-PC aligned brain and follow-up native T1 brain images. In parallel, diffusion MRI was corrected for motion and eddy-current distortion in FSL, then EPI susceptibility distortion was minimized by applying deformation maps generated from nonlinear co-registration between DWI b0 brain images and T1-weighted images at each time-point using ANTs (Advanced Normalization Tools). Subsequently, tensor reconstruction was performed in MRtrix3. Tensor and FLAIR images were then linearly co-registered to corresponding T1 AC-PC images at each timepoint.

#### 3. DTI data processing:

Diffusion weighted MRI data (dMRI) were pre-processed using tools provided by the software suites MRtrix3, FSL, and ANTs. Specifically, dMRI data were first denoised, then potential Gibbs-ringing artefacts were removed. The dMRI data were then corrected for bias field inhomogeneities using the ANTs N4 algorithm. a dMRI brain mask was then estimated using BET and used as an input, alongside the dMRI data, to eddy in order to correct for subject movement in the acquisition. Finally, phase distortion correction was applied using a non-linear registration method outlined below.

To correct for phase distortion within the dMRI data, first the brain was segmented from the corresponding T1w dataset, and a single b0 volume was extracted from the dMRI dataset. The T1w brain was then used as a mask to invert the contrast of the T1w image. A rigid-body registration was then performed on the inverted T1w image with the b0 volume as a target, which aligned the two images spatially. Non-linear registration was then performed using ANTs. The registration steps were comprised of a rigid body, then affine, and then SyN registration algorithm. The transformations and warps calculated from the non-linear registration steps were then applied to the entire dMRI dataset to correct for phase distortion artefacts.

**4. Identification of new acute lesions.**

Additional steps were therefore undertaken to identify and remove acute lesions that occurred during the study period.

Gadolinium-enhancing lesions that were detected at any time point were considered “new lesions” and excluded from the analysis of chronic lesion expansion for the next annual interval. However, gadolinium enhancement within active MS lesions usually does not persist beyond 2 months, after which newly formed T2 hyperintense lesions continue to shrink in size for another 3-5 months, reflecting resolution of edema and, potentially, tissue repair including remyelination [1]. Therefore, to accurately identify recent acute lesions, we further examined all lesions markedly (> 20%) shrinking between two consecutive time-points; and determined whether these lesions were newly formed at the previous time-point or already existing. All newly formed lesions at the previous time point were classified as 'acute lesions' and were only included in the chronic lesion analysis from the subsequent time-point onwards.

#### Supplementary figure legends.

Supplementary Fig. 1. Correlation between gradients of CLT enlargement obtained using manual vs IQ-MS$^{TM}$ AI-based lesion segmentation.

Supplementary Fig. 2.Violin plot showing Root Mean Square Error of cumulative CLT enlargement rate for the entire cohort.

Refferences.

1. **Rovira A**, **Auger C**, **Alonso J**. Magnetic resonance monitoring of lesion evolution in multiple sclerosis. *Ther Adv Neurol Disord*. 2013; **6**(5):298–310.

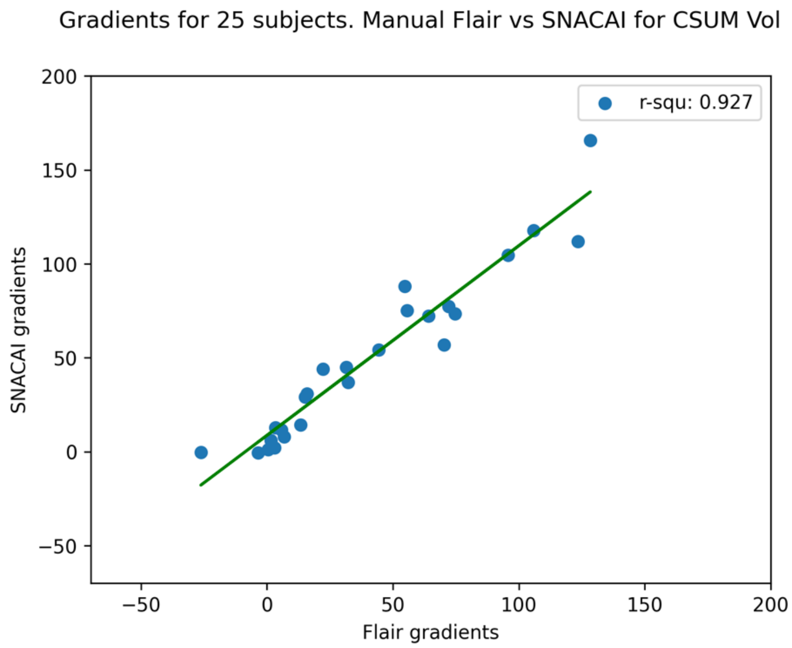
Gradients for 25 subjects. Manual Flair vs SNACAI for CSUM Vol
r-squ: 0.927
SNACAI gradients
Flair gradients
200
150
100
50
0
−50
−50
0
50
100
150
200

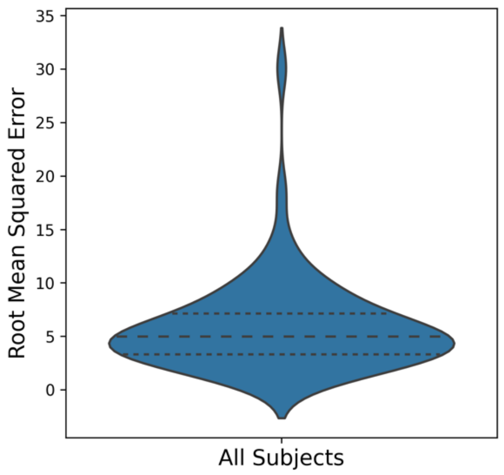
Root Mean Squared Error
All Subjects
35
30
25
20
15
10
5
0
